# Small Hands, Big Ideas: Exploring Nurturing Care Through Beatrice Alemagna’s “What is a Child?”

**DOI:** 10.1101/2025.01.04.25319990

**Authors:** Claudia Ravaldi, Alfredo Vannacci

## Abstract

This study examines the use of Beatrice Alemagna’s picture book "What is a Child?" as a tool for understanding and implementing nurturing care principles in perinatal healthcare settings. Through mixed-methods analysis of forum discussions among 42 perinatal professionals, including psychologists, midwives, physicians, nurses, pharmacists and early childhood educators, we explored how engagement with the book’s metaphorical and visual elements facilitated professional reflection and development. Qualitative analysis revealed four primary themes: the power of metaphors in conveying complex caregiving concepts, the importance of collective care and community support, understanding the temporality of care, and fostering transformative dialogue. Quantitative analysis demonstrated significant differences in thematic engagement across professional groups, with medical professionals emphasizing physical health (20.8%) and nutrition (32.1%), while psychologists focused more on early learning opportunities (28.3%) and responsive caregiving (21.0%). However, the picture book’s narrative framework encouraged professionals to transcend their domain-specific perspectives, fostering a more holistic appreciation of nurturing care principles. These findings suggest that carefully selected picture books can serve as sophisticated tools for professional development in perinatal care, particularly in translating abstract theoretical concepts into actionable insights while honoring different professional perspectives and expertise.

## 1. Introduction

The first thousand days of life, spanning from conception to age two, represent a critical period for child development. During this time, the interplay of experiences and environmental factors significantly influences brain development, learning, health, behavior, and future social relationships and economic productivity. The World Health Organization’s Nurturing Care Framework highlights the importance of this period, emphasizing that nurturing care can mitigate health inequities and promote optimal development. This framework identifies five interrelated components: good health, adequate nutrition, safety and security, opportunities for early learning, and responsive caregiving. Each component is essential and synergistic, contributing to the holistic development of the child. Good health is foundational, encompassing both preventive care and the management of childhood illnesses. For example, maternal health during pregnancy is crucial, as it directly affects the child’s health outcomes. Research indicates that maternal care mediates the effects of nutrition and responsive stimulation interventions on children’s growth, underscoring the multifactorial nature of growth and development, which includes caregiver-child interaction and maternal education (Brown et al. 2017). Moreover, adequate nutrition is vital, beginning with maternal nutrition and continuing through practices such as exclusive breastfeeding and appropriate complementary feeding. Studies have shown that responsive feeding practices are linked to better nutritional outcomes in children, particularly in low- and middle-income countries (Bentley, Wasser, and Creed Kanashiro 2011). Safety and security are also paramount, as children must be protected from violence, abuse, and environmental hazards. The quality of caregiving plays a significant role in ensuring safety; for instance, caregiver insightfulness can mitigate the risks associated with exposure to violence, thereby promoting resilience in children (S. Gray et al. 2015). Furthermore, opportunities for early learning emerge through daily interactions and experiences, which are essential for cognitive and social development. Research has demonstrated that children learn effectively through observation and imitation, particularly in nurturing environments where caregivers engage actively with them (Salalι et al. 2019). Responsive caregiving, defined as the ability to understand and respond to children’s cues and needs, serves as the bedrock for the other components of nurturing care. Evidence suggests that responsive caregiving is associated with improved socioemotional outcomes in children, reinforcing the need for interventions that enhance caregiver responsiveness (Scherer et al. 2019). Additionally, caregiver responses to children’s emotional states can significantly influence children’s behavioral outcomes, such as the development of overeating behaviors linked to emotional regulation (Ju 2024). The importance of caregiver sensitivity is further highlighted by studies indicating that it predicts how infants utilize caregivers for distress regulation, which is crucial for emotional development (Riddell et al. 2011). When children receive nurturing care across all five domains, they are better positioned to develop the cognitive, social, emotional, and behavioral capacities necessary for thriving throughout life. Conversely, a lack of nurturing care can lead to adverse developmental outcomes, with implications that may extend across generations. For instance, children raised in environments lacking responsive caregiving may face increased risks of maladaptive behaviors and emotional difficulties, particularly in contexts of adversity (Kerns et al. 2014).

Thus, the five dimensions of the Nurturing Care Framework offer an essential roadmap for supporting a child’s development and reducing health disparities. These interconnected components—good health, adequate nutrition, safety and security, opportunities for early learning, and responsive caregiving—function in synergy to create the conditions for optimal growth and well-being. Addressing these dimensions in a holistic manner is vital to mitigating risks and fostering resilience in the critical early years of life. However, bridging the gap between theoretical frameworks and practical application remains a significant challenge, requiring innovative strategies to translate these principles into everyday practices that effectively support both children and their caregivers (Ravaldi 2024). One of the primary challenges is the complexity of the nurturing care framework itself, which can be overwhelming for both professionals and families. While the scientific evidence supporting nurturing care is robust, bridging the gap between theory and application requires effective communication strategies. Healthcare providers and early childhood professionals often struggle to convey these concepts in a manner that is accessible and actionable, especially given the time constraints of clinical encounters and the diverse needs of families (Costa et al. 2022). This highlights the necessity for training programs that equip professionals with the skills to translate theoretical knowledge into everyday practices. Moreover, traditional teaching methods and clinical guidelines frequently overlook the emotional and relational dimensions of nurturing care. The use of technical language and prescriptive approaches can create barriers to understanding and engagement, particularly for families from diverse cultural and educational backgrounds (Black et al. 2019). Effective communication tools that facilitate meaningful dialogue about sensitive topics, such as responsive caregiving and emotional attunement, are essential for fostering engagement and understanding (Lucas, Richter, and Daelmans 2017). Sustained engagement from both professionals and families is crucial for the successful implementation of nurturing care. Information alone is insufficient; approaches that inspire reflection and motivate behavioral change are necessary, particularly in resource-constrained settings where competing demands can overwhelm families (Kodish et al. 2019). This underscores the importance of developing interventions that not only provide information but also foster emotional connections and support systems among caregivers (Bergmeier et al. 2020).

In response to these challenges, innovative yet accessible approaches are needed. Here we propose that picture books may emerge as a particularly promising tool, offering several unique advantages. In particular, they provide a universal language that transcends professional and cultural boundaries; through their combination of narrative and visual elements, they can make complex concepts tangible and relatable. Moreover, high-quality picture books can create a shared space for reflection and dialogue, enabling both professionals and families to explore nurturing care principles in ways that feel natural and non-threatening (Ravaldi 2025). Picture books also offer practical advantages in terms of implementation. They are portable, reusable, and require no special training or equipment to use effectively. They can be integrated into existing services and programs with minimal additional resources. Perhaps most importantly, they provide a medium through which professionals can model the very qualities - such as attention, responsiveness, and emotional engagement - that are central to nurturing care.

### 1.1 Aim of the study

While the theoretical foundations of the Nurturing Care Framework provide a comprehensive guide to promoting child development, the challenge lies in translating these principles into practical, actionable strategies that resonate with caregivers and professionals. Recognizing this gap, our study set out to explore how these theoretical components could be effectively integrated into the daily practices of perinatal health professionals. Specifically, we sought to examine the potential of Beatrice Alemagna’s *What is a Child?* (Alemagna 2016) as a tool to facilitate this translation. The primary aim of this study was twofold. First, it aimed to evaluate how health professionals in perinatal care — midwives, psychologists, doctors, nurses and early childhood educators — engage with and interpret the nurturing care principles when prompted by the themes and metaphors presented in *What is a Child?*. Second, it sought to collaboratively define the potential characteristics and applications of high-quality picture books as resources for professional development and caregiver education. Through a semi-structured, practice-oriented training module and guided discussions, participants were invited to reflect on the book’s poetic narratives and rich visual metaphors. These elements were designed not only to evoke deeper empathy and self-awareness but also to provide a shared framework for discussing the emotional and relational aspects of caregiving. By integrating participants’ reflections with expert commentary, the study aimed to uncover the transformative potential of picture books as tools for professional growth and the implementation of nurturing care practices.

## 2. Methods

### 2.1 Study Design

We performed a mixed method study investigating the role of picture books as tools for fostering professional reflection and implementing nurturing care principles in perinatal support contexts. In particular, we examined forum-based discussions among perinatal professionals, conducted as part of an interactive, practice-oriented training module included in a post-graduate specialization course on early childhood development and perinatal care (*I Primi Mille Giorni* – The First Thousand Days, offered by Florence University since 2023). The course aims to enhance professionals’ understanding and application of nurturing care principles, emphasizing both theoretical foundations and practical implementation. The forum discussions analyzed here were designed to serve as a dynamic and collaborative learning environment, where participants could engage deeply with the themes presented in several sources, including selected picture books. The discussions were initiated following a live presentation of the book and a general overview of the principles of nurturing care. This introductory session was deliberately kept broad, without providing detailed guidance on practical application, in order to allow participants the freedom to explore and interpret the themes independently. A more structured framework for nurturing care practices was introduced later, during the discussion of the results derived from the qualitative analysis of the forum contributions. To ensure confidentiality, all responses were analyzed anonymously, and data were securely stored and processed in compliance with ethical guidelines.

### 2.2 Theoretical framework

This analysis focuses on the collective reflections stemming from discussions on the picture book “Che cos’è un bambino” (Alemagna 2008) (ed. Topipittori, 2008), the Italian version of Beatrice Alemagna’s *"What is a Child?"* (Alemagna 2016) (ed. Tate Publishing, 2016), selected for its profound ability to evoke deep insights into caregiving through its poetic and metaphorical exploration of childhood (**Figure 1**). The book offers a nuanced portrayal of children as "small people" with "big ideas", emphasizing their individuality, agency, and emotional complexity. Its dual focus on the present completeness of children and their ongoing growth highlights the dynamic and evolving nature of nurturing care. This perspective encourages caregivers to remain attentive to both the immediate needs of the child and their developmental trajectories. Picture books, especially those of high artistic and narrative quality, have the unique ability to engage readers on multiple levels, intellectual, emotional, and relational, thus allowing, in keeping with the principles of relational care, to create a shared space for dialogue and emotional connection. This relational dynamic is critical in professional settings, where fostering empathy and collaboration can significantly enhance caregiving practices. Furthermore, picture books like *"What is a Child?"* transcend the dichotomy between explicit messaging and artistic integrity. While they address core caregiving themes, they do so in ways that invite interpretation and personal resonance, avoiding didacticism. This quality allows the text to act as an "emotional mirror", reflecting the caregiver’s internal experiences and facilitating deeper self-awareness. The selection of *What is a Child?* was guided by its universal accessibility and broad appeal, with compelling visuals and evocative text that engage diverse audiences, regardless of their familiarity with literature or theoretical frameworks (Nam 2023). This inclusivity makes the book a versatile and effective tool for fostering reflective practices in both professional and personal caregiving contexts, exemplifying the capacity of picture books to frame complex concepts through textual and visual elements, engaging both intellectual and emotional dimensions to provoke thought and exploration.

**Figure 1.**
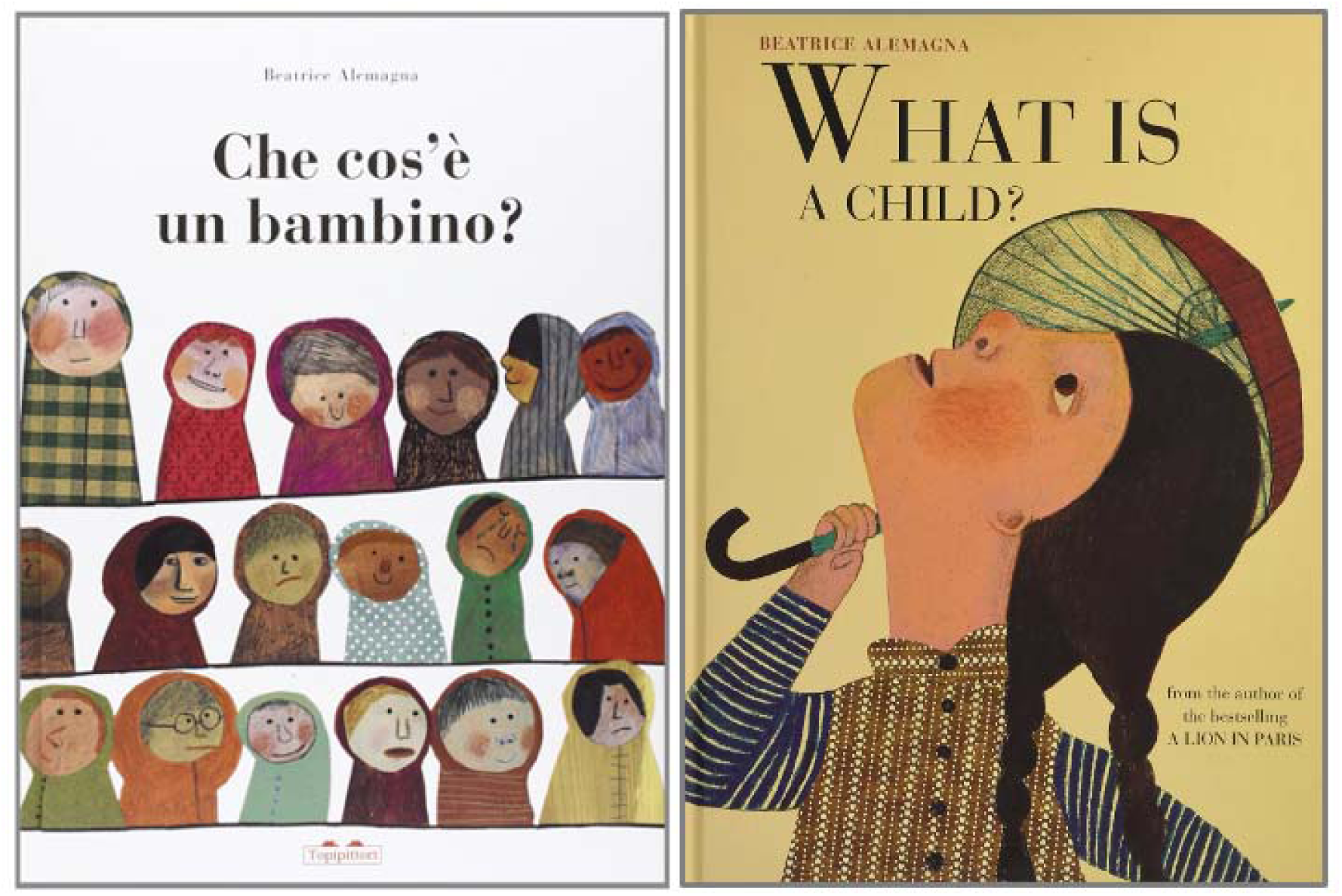
Covers of the Italian (left, Topipittori, 2008) and English (right, Tate Publishing, 2016) versions of the book.

### 2.3 Mixed Methods analysis

This study employed a mixed-methods approach, combining qualitative thematic analysis with quantitative evaluation, to comprehensively explore how the five domains of nurturing care, as defined by the World Health Organization (WHO)—good health, adequate nutrition, responsive caregiving, opportunities for early learning, and safety and security—were reflected in forum discussions. This approach was chosen to integrate the depth and nuance of qualitative insights with the comparability and rigor of quantitative analysis, ensuring a thorough understanding of the dataset and its alignment with nurturing care principles.

#### 2.3.1 Thematic analysis

The analysis began with the development of a framework for thematic classification. Keywords and phrases associated with each domain were curated based on foundational texts, perinatal care guidelines, and literature on early childhood development. To ensure comprehensive coverage, two AI tools were used to expand the keyword list, identifying synonyms and related terms to capture variations in language and context: ChatGPT o1 (OpenAI 2024) and Claude 3.5 Sonnet (Anthropic 2024). Forum responses were then scanned for the presence of these keywords, and each response was assigned to one or more themes based on keyword occurrence, as described in (Claudia Ravaldi et al. 2023). The process was repeated independently using both AI platforms to validate consistency, achieving 100% concordance across iterations.

#### 2.3.2 Quantitative analysis

Thematic scores were then calculated by counting the frequency of keywords for each response, and these scores were normalized by expressing keyword occurrences per 1,000 words to account for variations in response length. Aggregated scores for each theme were then analyzed to generate insights into the distribution and emphasis of the nurturing care domains. Two complementary metrics were used: proportional representation, which compared the total keyword occurrences for each theme to the overall keyword count, and normalized density, which highlighted the prevalence of thematic content across responses.Quantitative data analysis was conducted using Stata BE 18 (StataCorp) after anonymizing and importing the forum responses, which were downloaded from the Moodle LMS platform (moodle.org). Categorical data were reported as frequencies and percentages and compared using the chi-squared test, while continuous data were reported as mean values with standard deviations (SD) or medians [quartiles] and compared using t-tests or Kruskal-Wallis and Dunn tests for non-parametric comparisons, with statistical significance set at *p* < 0.05. Open-ended responses were coded and categorized using MAXQDA 2018, linking thematic categories to their corresponding nurturing care domains, and these quantified scores were integrated with the qualitative themes to identify patterns and variations. Data visualization, including proportional representations and normalized densities, was conducted using Tableau Desktop 2024.3 and Stata BE 18, offering a comprehensive view of the thematic emphasis within the dataset.

## 3. Results

Participants to the *I Primi Mille Giorni* course included a diverse group of 42 female perinatal professionals (median age 34 [29;40] ranging from 25 to 71), comprised of psychologists (17), midwives (14), physicians (2), nurses (2), pharmacists (3) and early childhood educators (4), who brought perspectives grounded in their respective fields of expertise. The forum threads were initiated by the moderators (CR and AV) with open-ended prompts encouraging participants to reflect on the concepts illustrated in Alemagna’s book and their relevance to nurturing care principles and their professional practice. These discussions generated a rich dataset comprising 68 posts, resulting in a total of 27,157 words that were analyzed qualitatively.

### 3.1 Thematic Insights from Forum Discussions

The qualitative analysis of forum discussions among perinatal care professionals unveiled four primary themes that align with the nurturing care principles, suggesting that the picture book may act as a bridge between reflective understanding and professional practice. These themes highlight the depth of engagement and the transformative impact of the text on caregiving perspectives. Below is an in-depth exploration of these themes with illustrative excerpts from the participants’ reflections.

#### 3.1.a The Power of Metaphors in Conveying Complex Ideas

The poetic and metaphorical language of *What is a Child?* emerged as a central feature in the forum discussions, with participants reflecting on how its vivid imagery and evocative style facilitated the understanding of complex caregiving concepts. The metaphors employed in the book were not merely illustrative but acted as bridges between abstract theoretical ideas and the lived experiences of caregiving. These reflections underscore the potential of metaphors to translate nuanced aspects of nurturing care into accessible and impactful tools for both professional and personal growth. Participants frequently emphasized how specific imagery in the book resonated deeply on both intellectual and emotional levels. The phrase “gentle eyes,” for example, stood out as a succinct yet profound encapsulation of responsive caregiving. One participant explained:

*"Alemagna writes, ‘children’s ideas are sometimes huge (…) they make their mouths open wide and say AH,’ but also, ‘children want to be listened to with wide-open eyes,’ and ‘to console them, gentle eyes are needed.’ These phrases reflect the creation of a climate of trust and listening, where the child feels welcomed and encouraged to express themselves."*

This metaphor was widely interpreted as a call to action for caregivers to embody a mindful and attentive presence. The participant reflecting on this metaphor connected it directly to their daily practice, emphasizing the importance of moving beyond routine caregiving tasks to prioritize relational and emotional dimensions. By fostering a space where children feel seen and heard, caregivers can cultivate a sense of safety and encouragement that supports both immediate and long-term developmental outcomes. Other metaphors elicited reflections on the emotional and physical safety central to nurturing care. One participant explained:

*"We also find attention to children’s physical environments in the phrase ‘children have small things, just like them: a small bed, small colorful books, a small umbrella,’ which reflects adults’ consideration for children and their need for objects suited to their size."*

This observation highlights the duality of caregiving as both a practical and emotional endeavor. While the original metaphor stems from a physical representation of comfort, participants connected it to broader concepts of safety and nurturing. This emphasis on creating supportive environments is particularly relevant in contexts of vulnerability, where physical and emotional security are paramount. The forum also revealed that metaphors acted as “emotional mirrors,” prompting participants to introspect about their own caregiving experiences. For example, one participant noted:

*"Children are like sponges. They absorb everything: nervousness, bad ideas, the fears of others. They seem to forget, but then it resurfaces in their school bags, under the covers, or in front of a book."*

This reflection connects directly to the principles of responsive caregiving and self-regulation, demonstrating how metaphors can facilitate a deeper awareness of the caregiver’s role not just in action but in emotional presence.

Metaphors therefore transcended their textual origins to become tools for professional reflection. By linking abstract concepts to tangible images and personal experiences, the book helped participants reframe their understanding of nurturing care. This capacity to evoke reflection through metaphor was particularly significant in enabling professionals to connect theoretical principles with their own caregiving practices, thereby enhancing their ability to provide holistic and empathetic care.

#### 3.1.b Collective Care and the Role of the Community

The forum discussions underscored the importance of collective care, revealing how professionals interpreted caregiving as a shared responsibility. This theme aligns with the principle that "it takes a village to raise a child," a concept central to nurturing care frameworks (C. Ravaldi 2024). Participants reflected on how the illustrations and text of *What is a Child?* highlighted the interconnectedness of caregivers and their communities, prompting a re-evaluation of caregiving as a collaborative endeavor rather than an individual task. One participant wrote:

*"Parents also need support in facing parenthood, as can happen with those children described in the book as ‘difficult, unbearable, (…) who break plates, bowls, and everything else.’ Parents too need a caregiver to support them through responsive care in this journey, so that they, in turn, can do the same for the child."*

This insight illustrates a paradigm shift for many professionals, who often work within systems that emphasize individual responsibility in caregiving. In this case, the book acted as a catalyst for recognizing the broader relational networks that contribute to a child’s development, including extended family, peers, and community support structures. The participant’s reflection also emphasizes the duality of caregiving, where supporting the parent is equally essential for fostering the child’s well-being. This perspective is particularly relevant in the context of perinatal care, where the burden on primary caregivers—often mothers—is disproportionately high, leading to burnout and reduced caregiving efficacy. By highlighting the need for caregivers to be cared for themselves, the discussion reframes caregiving as a reciprocal and collective endeavor. Another participant expanded on this notion:

*"A child who grows without us noticing gives me the sense that someone sufficiently competent was there to ensure their growth wasn’t made difficult or burdensome."*

This reflection underscores the critical role of caregivers in providing consistent and competent support, enabling children to thrive without undue stress or obstacles. Participants emphasized that this form of caregiving often requires a network of relationships, including family members, educators, and peers, working collaboratively to support the child. This insight aligns with the nurturing care framework’s emphasis on creating environments where children feel secure and supported in their development. Discussions revealed that professional settings often neglect the broader relational networks that influence caregiving outcomes, focusing instead on the immediate caregiver-child dyad. Participants noted that fostering a collaborative approach, involving extended family and community resources, could significantly enhance caregiving practices, particularly in resource-constrained environments. Such reflections highlight the importance of creating not only a direct bond between caregiver and child but also a wider ecosystem of care, ensuring that children receive the comprehensive support necessary for their growth and well-being.

The relational focus of *What is a Child?* also invited introspection about participants’ roles within these caregiving ecosystems. Several professionals commented on the need to model collaborative practices in their own work, creating environments where both families and caregivers feel supported. Discussions frequently returned to the metaphorical "village," which participants viewed as essential not only for children but for caregivers themselves. One professional shared:

*"It is a well-established truth that ‘to raise a child, it takes a village.’ Children change, just as the world of a woman, a man, a couple, or a caregiver is literally turned upside down as they face the new reality of parenthood. They should be guaranteed the opportunity to invest their efforts in a way that is sustainable for them."*

This reflection highlights a critical dimension of collective care: the well-being of the caregivers themselves. Participants discussed how their own professional networks—or lack thereof—shaped their capacity to deliver nurturing care. The forum thus became a space for mutual support, as participants shared strategies for building and sustaining their own "villages."

Finally, the discussions touched on systemic barriers to collective caregiving, such as cultural expectations, economic constraints, and policy gaps. While the book inspired idealistic visions of interconnected caregiving, participants acknowledged the practical challenges of implementing these ideals. As one participant noted:

*"It is also necessary to promote good healthcare and efficient access to the services needed. The book mentions ’children with glasses, in wheelchairs, with braces on their teeth,’ highlighting how not everyone has the same opportunities for adequate care, creating significant inequalities."*

These discussions around collective care revealed the book’s transformative potential to inspire reflections on caregiving networks, both personal and professional. By reframing caregiving as a shared responsibility, the book challenged participants to envision more inclusive and supportive systems, while also acknowledging the barriers that remain. This theme underscores the importance of integrating relational and community-based approaches into the nurturing care framework, highlighting both opportunities and challenges for systemic change.

#### 3.1.c Understanding the Temporality of Care

Forum discussions revealed deep reflections on the temporality of care, a concept explored in What is a Child? through its dual emphasis on children as "complete in the moment" while simultaneously "constantly evolving." This temporal duality emerged as particularly significant for understanding how nurturing care principles adapt across developmental stages. The book’s treatment of temporality prompted participants to reflect on three key dimensions: individual developmental rhythms, intergenerational impact and the dynamic nature of caregiving. Some participants emphasized how the text encouraged recognition of each child’s unique developmental pace:

*"The importance of time in child development and growth: each is a small human being with their own needs and pace, unique and different, which caregivers must respect and support without forcing, while constantly providing guidance and support.”*

This reflection emphasizes the book’s ability to prompt consideration of individualized care approaches that honor each child’s developmental trajectory. The text also sparked consideration of how present care shapes future outcomes. As one participant observed:

*"Children who decide not to grow up will never grow. They will have a mystery within them. Even as adults they will be moved by small things: a ray of sunshine or a snowflake.”*

This observation highlights how early experiences influence lifelong emotional patterns, emphasizing the long-term implications of nurturing care. Finally, participants also reflected on how caregiving itself evolves over time:

*"A child grows without us even noticing… slowly and silently, their body lengthens. A child is not a child forever*.

*One day they will change.”*

This insight underscores how caregiving must adapt responsively to children’s changing needs while maintaining consistency in core nurturing principles. These discussions revealed how the book’s treatment of temporality helped professionals conceptualize nurturing care as both anchored in the present moment and oriented toward future development - a perspective that enriches understanding of how to implement responsive care across the developmental continuum.

#### 3.1.d Fostering Transformative Dialogue

Forum discussions highlighted how *What is a Child?* encouraged parents and caregivers to reflect on their own emotions, past experiences, and caregiving practices. The evocative text and illustrations fostered a process of self-awareness, helping professionals and families alike to better understand the relational dynamics at play in caregiving. One participant shared:

*"Children might pretend not to notice ’the parent’s tears in the shower,’ but they can perceive distress, sensing that something is wrong. It’s important to guide and support them in their growth, which is not only physical but also psychological."*

Another noted:

*"Browsing through the pages of this book, parents are confronted with situations they experienced and that their child is living in the present. This process stimulates awareness, helping them see their child for who they are, without projecting dreams and desires that belong to their own childhood."*

Participants emphasized the need for empathy and emotional attunement, where the parent or caregiver becomes an active participant in the child’s developmental journey. The book’s narrative depth facilitated this process by enabling parents to revisit their childhoods and acknowledge how their past influences their caregiving. One professional reflected:

*"The greatest value of this book is its ability to stimulate, on a preconscious level, a reworking of one’s own past. This helps the parent to emotionally attune to the child, creating a connection that makes caregiving more rewarding and fulfilling."*

These reflections illustrate how storytelling can foster a deeper connection between parents and children, bridging gaps in understanding and creating spaces for mutual growth and healing. The book served as both a mirror and a guide, prompting adults to engage in a transformative process of self-discovery and emotional resonance with the children in their care.

### 3.2 Quantitative evaluation of Nurturing Care Domains

The quantitative analysis of the forum responses revealed a rich distribution of the five themes of nurturing care across the dataset. **Table 1** provides a summary of the occurrences, proportional representation, and density of each theme per 1000 words, offering insights into the thematic emphasis in professional reflections.

**Table 1:** Distribution and Density of Nurturing Care Themes in Forum Discussions.

| Theme | Occurrences | Percentage | Density |
| --- | --- | --- | --- |
| Promoting Good Health | 292 | 24,35% | 10,75 |
| Ensuring Adequate Nutrition | 193 | 16,10% | 7,11 |
| Providing Safety and Security | 240 | 20,02% | 8,84 |
| Responsive Caregiving | 289 | 24,10% | 10,64 |
| Opportunities for Early Learning | 185 | 15,43% | 6,81 |
| Total | 1199 | 100 | 44,15 |

**Figure 2** illustrates the percentage distribution of themes across the dataset, highlighting the relative importance of each domain. The data indicates a nearly equal emphasis on Promoting Good Health and Responsive Caregiving, which collectively account for almost half of the thematic content.

**Figure 2:**
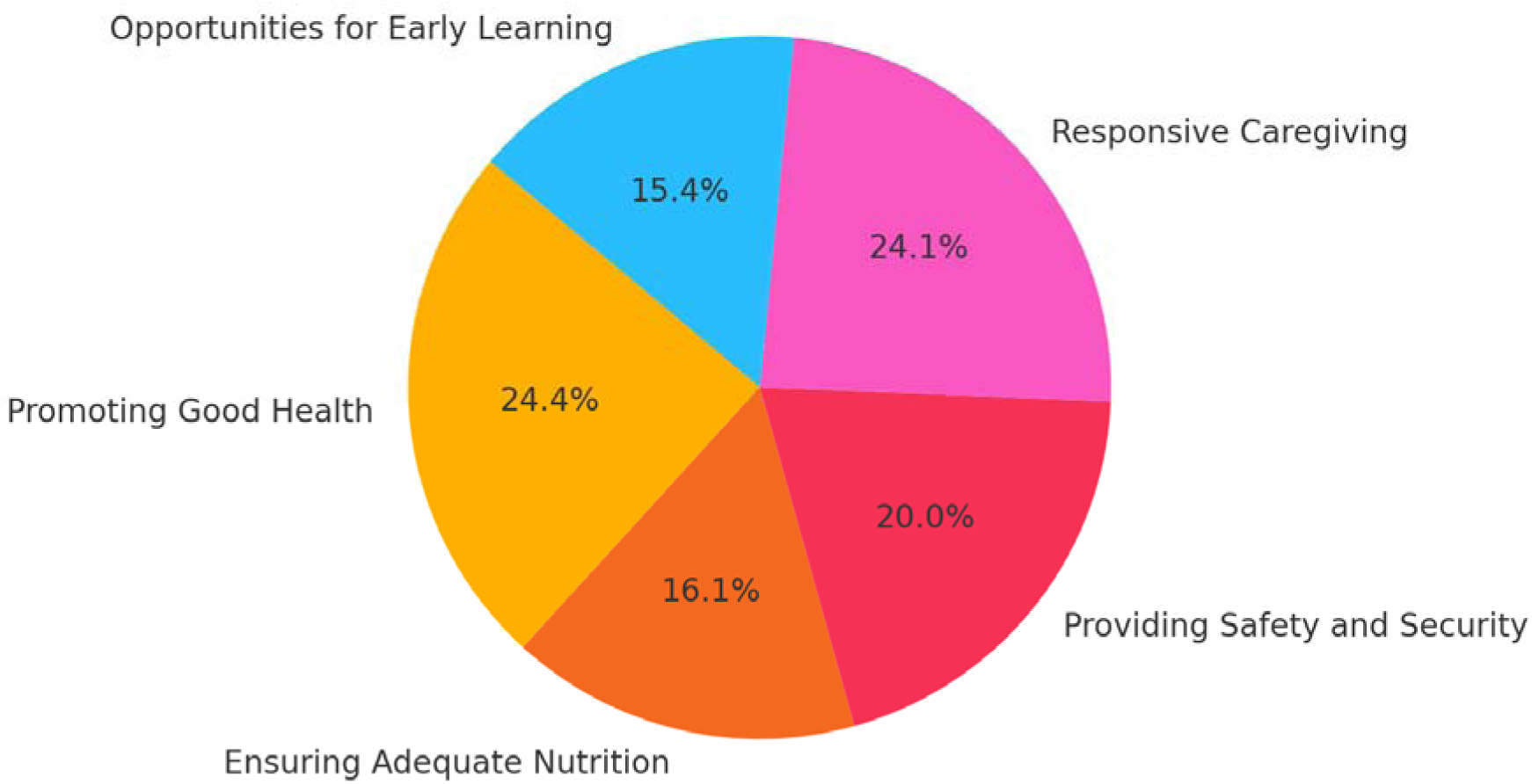
Percentage Distribution of Nurturing Care Themes in Forum Discussions

**Figure 3** presents the density of thematic keywords, normalized to occurrences per 1,000 words. The highest densities are observed for Promoting Good Health (10.75) and Responsive Caregiving (10.64).

**Figure 3:**
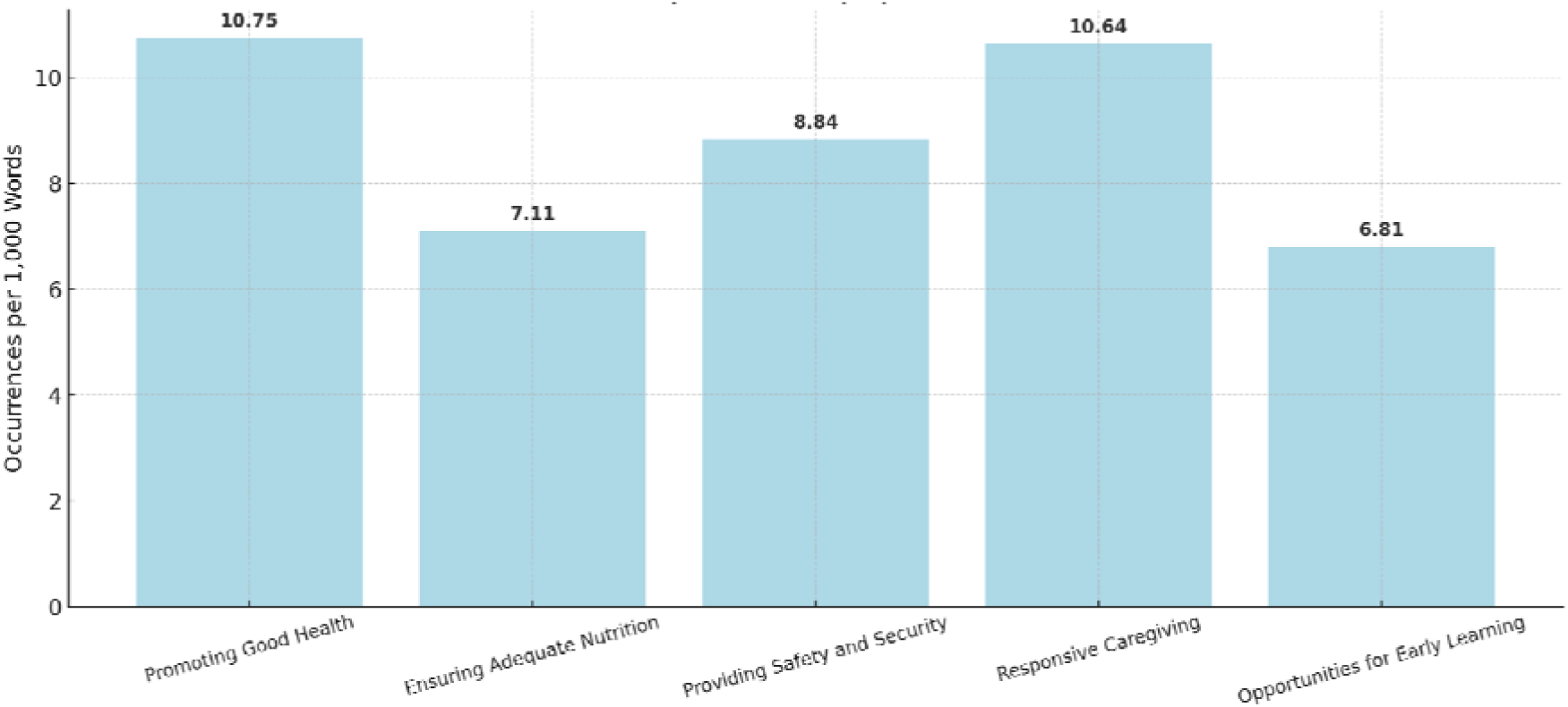
Keyword Density (per 1000 words) of Nurturing Care Themes in Forum Discussions.

**Figure 4** explores how thematic priorities vary across professional groups. Medical professionals (comprising physicians, pharmacists, and nurses) were grouped together due to their shared clinical approach, which differed significantly from that of psychologists and early childhood educators. The most notable differences were observed between the medical group and psychologists, particularly in themes of Ensuring Adequate Nutrition, Opportunities for Early Learning, and Promoting Good Health. Medical professionals prioritized Promoting Good Health and Ensuring Adequate Nutrition, reflecting their clinical focus on tangible health outcomes. In contrast, psychologists placed greater emphasis on Providing Safety and Security and Opportunities for Early Learning, aligning with their expertise in relational and developmental aspects. Early Childhood Educators demonstrated a balanced focus on Responsive Caregiving and Opportunities for Early Learning, emphasizing both relational and cognitive development.

**Figure 4:**
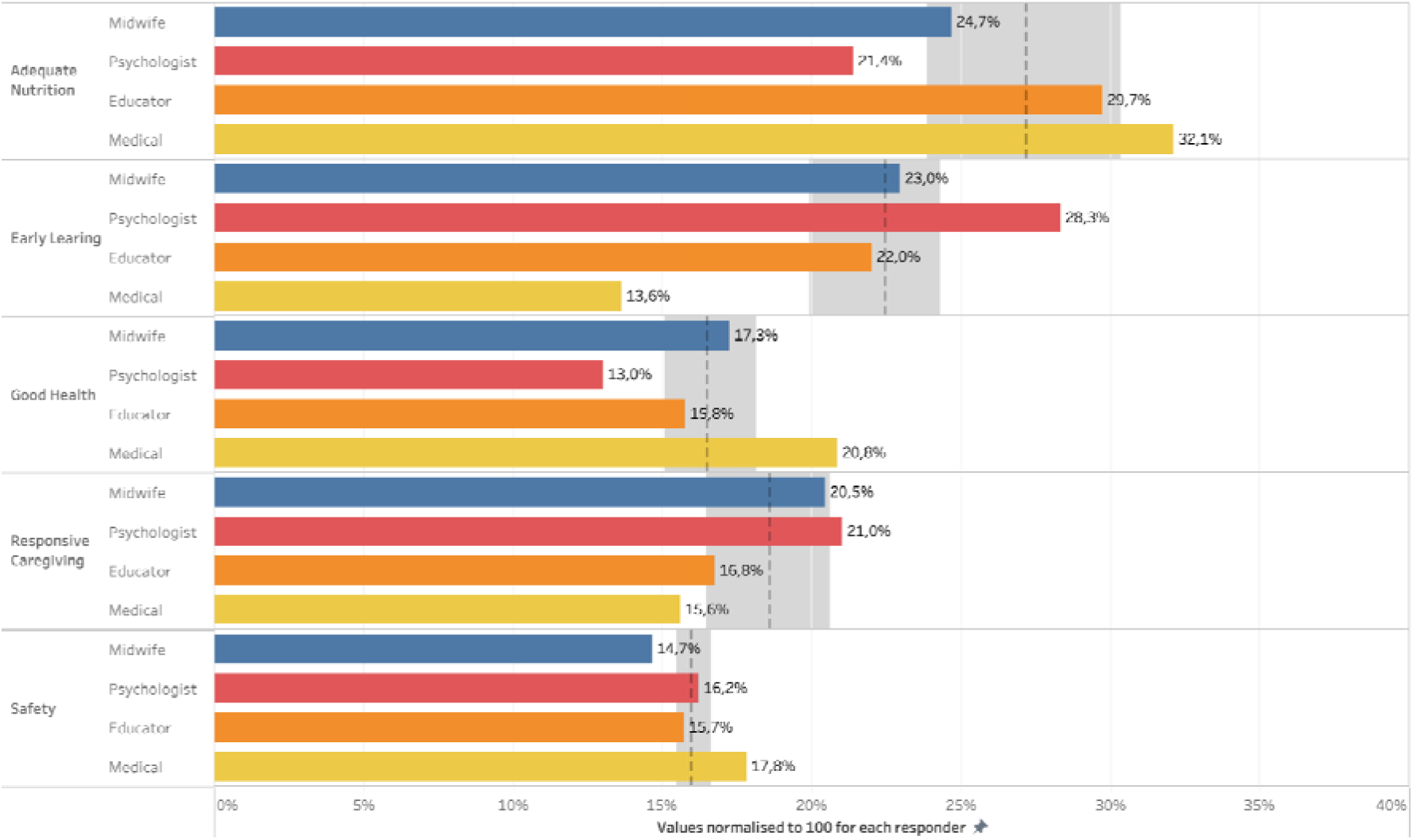
Relevance of nurturing care domains according to professional role. * p<0.01 vs Psychologists ; # p<0.05 vs Psychologists (Dunn test). Dotted lines show median values, shaded areas show quartiles. "Medical" category includes doctors, nurses and pharmacists.

## 4. Discussion

The analysis of forum discussions revealed a complex interplay between metaphorical understanding and professional expertise in engaging with nurturing care principles. The quantitative distribution of themes shows that "Promoting Good Health" (24.4%) and "Responsive Caregiving" (24.1%) emerged as the most frequently referenced domains, followed by "Providing Safety and Security" (20.0%). This distribution suggests that participants found the picture book particularly effective in eliciting reflections on these aspects of nurturing care. When examining keyword density, a similar pattern emerges, with "Promoting Good Health" and "Responsive Caregiving" showing the highest density of thematic references (10.8 and 10.6 occurrences per 1,000 words, respectively). This consistency between proportional representation and keyword density suggests that these themes were not only frequently mentioned but also discussed in depth, indicating that the picture book successfully facilitated engagement with core nurturing care concepts.

Professional background significantly influenced how participants engaged with different domains of nurturing care. Medically-oriented professionals showed a marked preference for discussing more “practical” themes, such as nutrition (32.1%) and physical health (20.8%), while psychologists emphasized early learning opportunities (28.3%) and responsive caregiving (21.0%). It is in fact known that different professional backgrounds shape the interpretation and discussion of themes related to care, such as practical health considerations versus emotional support, thereby reinforcing the notion that professional training and experience shape the interpretation of narrative materials (Tonini et al. 2021).

Notably, the metaphorical language of the picture book appeared to facilitate cross-disciplinary understanding. For instance, the metaphor of children as "sponges" allowed both medical professionals and psychologists to broaden their typical professional focus, considering more holistic implications for child development. This highlights the potential of picture books as tools for fostering interprofessional dialogue and understanding. Moreover, the integration of qualitative and quantitative data revealed how the narrative framework encouraged professionals to transcend their domain-specific perspectives, fostering a holistic appreciation of nurturing care. These findings align with existing literature on the role of humanities in healthcare education, which underscores the value of narrative and metaphor in promoting empathy and collaborative thinking (Reuter, Santos, and Ramos 2018). Such insights suggest that carefully selected picture books can effectively support the development of integrated approaches to perinatal care.

### 4.1 Picture Books as Tools for Professional Development

Our analysis reveals how picture books can serve as sophisticated tools for professional development in perinatal care, particularly in translating abstract theoretical concepts into actionable insights. The effectiveness of "What is a Child?" as a professional development tool manifested in three key dimensions: metaphorical understanding, visual-narrative engagement, and collaborative meaning-making.

#### Metaphorical understanding

The metaphorical language employed in "What is a Child?" serves as a bridge between theoretical knowledge and practical application. Research indicates that metaphor plays a crucial role in professional learning, particularly in fields such as healthcare where complex concepts must be understood and applied in real-world contexts. Several authors emphasize the importance of narrative techniques in enhancing understanding and engagement among learners, suggesting that metaphorical imagery can facilitate deeper cognitive connections. Research indicates that narrative techniques can significantly improve learners’ engagement and comprehension in educational contexts, particularly in health-related fields (Carrington, Allen, and Osmolowski 2007). Additionally, studies have shown that narratives in health promotion can enhance communication of knowledge and behavioral modeling, thereby fostering engagement and understanding (Kreuter et al. 2007). Building on this, the forum discussions surrounding the book revealed that metaphors like "gentle eyes" and children as "sponges" not only fostered personal reflection but also transformed theoretical concepts into actionable insights. This reflects the broader educational potential of metaphorical understanding, as emphasized by several authors, who highlight how structured narratives and symbolic imagery can facilitate the transfer of abstract knowledge into practical contexts (Romanchuk et al. 2023). Such approaches also resonate with findings on the effectiveness of narrative-based learning in promoting ethical understanding, bridging the gap between theory and practice in professional development contexts (Mawasi et al. 2022).

#### Visual-narrative engagement

The combination of visual and textual elements in "What is a Child?" created multiple entry points for professional learning. The quantitative analysis shows that different professional groups engaged preferentially with different aspects of the narrative—medical professionals focusing more on physical health representations (20.8% of their responses) while psychologists emphasized relational elements (21.0% of responses). However, the visual elements often prompted professionals to expand beyond their typical domain focus, supporting a more holistic understanding. Research indicates that visual literacy plays a crucial role in professional development, enabling practitioners to interpret and utilize visual information effectively in their practice (Zayeb, Aleidan, and Ali 2024). The book’s visual metaphors particularly supported engagement with complex concepts around safety and security (20.0% of total themes). As one participant observed: "The illustrations of ’small things for small people’ helped me understand environmental safety not just as physical protection but as emotional containment." This suggests that visual narratives can enhance understanding of abstract caregiving concepts, reinforcing the effectiveness of visual metaphors in professional learning environments.

#### Collaborative meaning-making

The forum discussions demonstrated how shared engagement with picture books can foster interprofessional dialogue and learning. The analysis shows that when discussing the same visual or textual elements, different professional groups often contributed complementary perspectives, enriching collective understanding. This aligns with research on the value of shared reading experiences in professional development, which highlights the importance of collaborative learning in healthcare settings (Jentoft 2021). Expert commentary from moderators often highlighted these moments of cross-professional insight. For example, when different professionals discussed the "sponge" metaphor, their diverse perspectives combined to create a more comprehensive understanding of child development. This suggests that picture books can serve as effective platforms for interprofessional education, facilitating dialogue and collaborative meaning-making among various healthcare professionals (Zaher et al. 2022).

### 4.2 Professional Identity and Domain-Specific Engagement

The quantitative analysis of how different professional groups engaged with nurturing care domains reveals distinct patterns of emphasis that reflect professional identity and training, while also suggesting opportunities for expanding professional perspectives through narrative engagement.

Our analysis revealed significant differences in how professional groups prioritized various aspects of nurturing care. Medical professionals showed a marked preference for nutrition and physical health, significantly higher than other groups. This emphasis aligns with traditional medical education’s focus on physical health outcomes (Frenk et al. 2010). In contrast, psychologists devoted more attention to early learning opportunities and responsive caregiving, reflecting their training in developmental and relational aspects of care (Zaher et al. 2022).

Midwives demonstrated a more balanced engagement across nurturing care domains, with relatively even distribution among health, nutrition, and responsive caregiving. This pattern suggests that midwifery training may incorporate a more holistic approach to perinatal care, emphasizing the interconnectedness of physical, emotional, and relational health. Educational models in midwifery often prioritize reflective practices and continuity of care, aligning with holistic, women-centered philosophies (Yanti et al. 2015). Beyond formal education, professional experiences further refine midwives’ approaches as well, enabling them to adapt to diverse perinatal scenarios and develop comprehensive care strategies that address the full spectrum of maternal and neonatal needs (Hollander et al. 2019). For instance, we recently highlighted that while only 33.4% of Italian midwives had received formal training in perinatal loss care, their practical exposure often allowed them to effectively navigate complex emotional and clinical situations (Ravaldi et al. 2022a), although this exposure also places them at significant risk of burnout and post-traumatic stress (Ravaldi et al. 2022b). In contrast, early childhood educators displayed a pronounced focus on connecting nurturing care principles to practical learning opportunities (22.0%), while maintaining attention to physical care needs. Their emphasis reflects a specific grounding in developmental and educational frameworks, highlighting the importance of interdisciplinary approaches in fostering holistic caregiving strategies.

Despite these professional differences, the forum discussions revealed how picture book engagement encouraged professionals to venture beyond their typical domains. For example, when discussing some metaphors of the book, medically-oriented professionals expanded their consideration of health to include emotional well-being, while psychologists incorporated physical health aspects into their reflections on responsive caregiving. While some differences were significant, qualitative analysis showed integration of multiple perspectives, suggesting that such engagement with narrative materials may support a holistic understanding.

The patterns observed in our analysis suggest that while professional identity strongly influences how caregiving is understood and discussed, carefully selected narrative materials can support more integrated approaches to nurturing care.

### 4.3 Theoretical and Practical Implications

The integration of qualitative and quantitative findings from this study contributes to both theoretical understanding and practical applications in professional development for perinatal care. These implications span several key areas that warrant detailed discussion.

This study extends current understanding of how narrative tools can support professional development in healthcare settings. While previous research has primarily focused on traditional literary texts as tools for narrative medicine (Milota, Van Thiel, and Van Delden 2019), our findings demonstrate that picture books offer unique advantages for professional learning. The combination of visual and textual elements appears to facilitate engagement with complex caregiving concepts, particularly for professionals working in perinatal care. This supports emerging theories about multimodal learning in healthcare education, where storytelling and reflective practices enhance communication, empathy, and critical thinking (Liao and Wang 2020). Our results also contribute to understanding how metaphorical thinking supports professional development. The quantitative analysis showed that metaphor-rich passages generated the highest keyword density, while qualitative analysis revealed how these metaphors facilitated the translation of theoretical concepts into practice. This extends current theory on metaphor in professional learning, demonstrating its utility in multiple domains. Metaphors serve as a powerful tool for constructing new knowledge within professional environments. They enable professionals to uncover tacit assumptions, articulate complex experiences, and envision new possibilities for their practice (Daley 2001). In healthcare, metaphorical thinking has been shown to clarify unwritten assumptions and motivators that shape professional behavior, making it a valuable strategy for understanding and addressing complexities in practice (Aita et al. 2003).

The findings suggest several practical applications for professional education in perinatal care. First, narrative tools like picture books can serve as effective interprofessional education tools. By offering shared storytelling frameworks, they enhance collaboration and communication among professionals while fostering deeper engagement with patient-centered care (Gray 2009). Second, the use of storytelling as an educational tool has been shown to improve critical skills like empathy, reflective thinking, and communication, making it a valuable component of healthcare training programs (Langer and Ribarich 2008).

For successful implementation, criteria like the relevance of the narrative, integration into broader training models, and the role of facilitation are crucial. Programs such as those described in surgical and interprofessional education demonstrate the ability of storytelling to strengthen professional identity, enhance teamwork, and translate theoretical concepts into practice (Scott-Conner and Agarwal 2022). These approaches not only improve professional competencies but also promote more humanistic healthcare delivery.

### 4.4 Limitations and Future Directions

Our study strongly suggests the value of picture books in professional development for perinatal care, while also highlighting important considerations for interpreting and applying these findings. Our analysis draws on responses from 42 female perinatal professionals, including psychologists, midwives, physicians, nurses, pharmacists and early childhood educators, all practicing in Italy. While this represents a diverse range of professional backgrounds within perinatal care, the demographic homogeneity of the participant group indicates the need for caution in generalizing across more diverse populations and cultural contexts.

Additionally, though the forum format encouraged rich discussion and reflection, producing over 27,000 words of qualitative data, written responses may not fully capture the nuanced ways professionals engage with picture books in practice. The asynchronous nature of forum discussions, while allowing for careful consideration and detailed responses, also lacks the immediacy of real-time engagement and non-verbal cues that might emerge in face-to-face professional development settings. Moreover, our focus on written reflection means we cannot observe how professionals might actively use these narrative tools in their practice with families and colleagues.

While "What is a Child?" proved particularly effective for exploring nurturing care principles, it represents one carefully selected example within the broader landscape of children’s literature. Our research group is currently conducting systematic studies to identify and validate the specific characteristics that make picture books effective tools for professional development and psychological support. This ongoing work aims to establish evidence-based criteria for selecting and implementing narrative materials in healthcare settings. Other high-quality picture books might offer different entry points for professional reflection and development, suggesting the value of expanding this line of inquiry.

The quantitative analysis revealed intriguing differences in engagement patterns across professional groups, but the mechanisms underlying these variations merit further investigation. Future research could enrich our understanding through direct observational studies of picture book use in training settings, longitudinal assessment of impact on professional practice, and cross-cultural studies of narrative engagement. Studies comparing different narrative tools could illuminate which features most effectively support professional development, while intervention studies could examine facilitation strategies to bridge professional differences. Additional research examining implementation across diverse healthcare settings would provide practical guidance for expanding this approach. These investigations would contribute to our evolving understanding of how narrative tools can support healthcare professionals in providing more holistic and responsive care, while maintaining appropriate empirical caution about the scope and generalizability of current findings.

## 5. Conclusion

This study highlights how picture books can serve as sophisticated tools for professional development in perinatal care, particularly in translating the abstract principles of nurturing care into actionable insights. Through mixed-methods analysis of forum discussions centered on Beatrice Alemagna’s "What is a Child?", we have shown how carefully selected picture books can create shared spaces for professional reflection and development while honoring different professional perspectives and expertise.

The integration of qualitative and quantitative findings reveals that picture books support professional development through multiple mechanisms. The metaphorical language provides accessible entry points for complex caregiving concepts, while visual elements create opportunities for multiple interpretations and shared understanding. The balanced engagement across nurturing care domains present in our analysis suggests that appropriate picture books can support comprehensive understanding of holistic care frameworks while respecting professional expertise.

Particularly noteworthy is how picture books appear to facilitate interprofessional dialogue and learning. While our analysis revealed significant differences in how professional groups prioritize various aspects of nurturing care, the shared engagement with narrative materials encouraged professionals to venture beyond their typical domains of expertise. This suggests that picture books may also serve as tools for promoting more integrated approaches to perinatal care.

These findings have important implications for professional education and practice development in healthcare settings. They suggest that picture books, when thoughtfully selected and skillfully facilitated, can support the development of more nuanced and holistic approaches to care while maintaining the valuable perspectives that different professional backgrounds bring to perinatal care.

As healthcare continues to evolve toward more integrated and person-centered approaches, tools that support professionals in understanding and implementing complex care frameworks become increasingly important. Picture books emerge from this study as promising resources for this purpose, offering accessible yet sophisticated means of engaging with the multifaceted nature of nurturing care. Future research exploring these possibilities across different healthcare contexts and cultural settings will further enhance our understanding of how narrative tools can support professional development and, ultimately, improve care for families during the critical perinatal period.

## Data Availability

All data produced in the present study are available upon reasonable request to the authors

## Acknowledgements

We wish to express our gratitude to all the participants of Florence University “*I Primi Mille Giorni”* course 2024-25, who contributed to the forum discussions and participated in the in-class dialogues that inspired the reflections elaborated in this work. While their contributions do not qualify them for authorship, we deeply appreciate their engagement with the themes presented in this study: Arianna Balestracci, Giada Barbirato, Elena Baritello, Simona Bensi, Elisa Bertilorenzi, Francesca Casartelli, Chiara Castracani, Marina Ceragioli, Cecilia Cirilli, Valentina D’Alesio, Giulia Fava, Anisia Fazzi, Irene Graziani, Marcella Marcone Terzago, Nausicaa Martino, Chiara Mauri, Federica Minelli, Albina Mingaj, Valentina Modolo, Tosca Mori, Miriam Morici, Laura Mosconi, Alice Pasquini, Noemi Passalacqua, Chiara Poli, Anna Pomes, Chiara Rossini, Althea Iris Salerno, Valentina Scarpello, Ambra Sciascia, Maria Selene Tarascio, Silvia Zanette, Alessandra Zani, and Laura Zanoni.

## 6. Statements and Declarations

### Contributors

CR (conceptualisation; investigation; writing—original draft preparation; review and editing: project administration); AV (conceptualisation; writing—original draft preparation; investigation; data management and analysis; review and editing). AV is the guarantor.

### Ethical statements

This study analyzed anonymised contributions from a professional forum. While no personal data were collected, participants consented to the use of their forum posts for research purposes, as explicitly mentioned in the forum’s participation guidelines. Additionally, all participants are acknowledged in the manuscript for their valuable contributions. The Ethics Committee for Research of the University of Florence does not require formal approval for studies of this nature, as they do not involve the collection of sensitive personal data or direct interaction with participants beyond the scope of their original forum activity.

### Funding

The study was not funded; no researcher received grants, salary or reimbursements for the realisation of the study.

### Competing interests

None declared.

### Data availability statement

Raw data and forum posts are available upon reasonable request.

### Patient and Public Involvement

Patients and the public were not involved in this research study

## Notes

### Competing Interest Statement

The authors have declared no competing interest.

### Funding Statement

This study did not receive any funding

### Author Declarations

This study analyzed anonymized contributions from a professional forum. While no personal data were collected, participants consented to the use of their forum posts for research purposes, as explicitly mentioned in the forum's participation guidelines. Additionally, all participants are acknowledged in the manuscript for their valuable contributions. The Ethics Committee for Research of the University of Florence does not require formal approval for studies of this nature, as they do not involve the collection of sensitive personal data or direct interaction with participants beyond the scope of their original forum activity.

